# Privacy Preserving Federated Gradient Boosting for Single Record Patient Devices

**DOI:** 10.64898/2026.02.10.26345891

**Authors:** Bernhard Specht, Orhan Ermis, Samaher Garbaya, Reinhard Schneider, Ricardo Chavarriaga, Djamel Khadraoui, Zied Tayeb

## Abstract

Learning from data on personal devices requires methods that accommodate limited local records, intermittent availability and privacy constraints. Here we present PrivateBoost, a federated gradient boosting framework that allows clients to contribute individual records without fitting local models or coordinating with other clients. Each device distributes cryptographic shares of its contribution to a fixed set of shareholders, and an aggregator reconstructs their sums for training. Simulations on four medical datasets containing up to 70,692 records show predictive performance comparable to matched centralised training and resilience to simulated client dropout. Per-client communication ranges from approximately 0.6 to 4.2 megabytes in loopback measurements for models with 15 trees of maximum depth three. In a physical deployment, four consumer smartphones and tablets, each holding a batch of records, complete three training sessions unattended during operating-system-managed background execution. Median share-generation times range from 0.3 to 1.0 milliseconds per round. These results demonstrate single-record learning in simulation and show that mobile participation can fit within brief background execution windows, without requiring users to keep an application open.

## Introduction

Developing medical prediction models requires access to clinical data that are often scattered across separate systems [1]. Bringing these records together raises questions about consent, privacy and responsibility for their use [2]. Federated learning allows participants to train a shared model by exchanging locally computed updates while retaining the underlying records [3]. In health care, it has been applied to collaborative brain tumour segmentation [4] and prediction of oxygen support requirements for patients with COVID-19 [5]. These applications demonstrate collaboration across institutions; participation from patients’ personal devices introduces different practical requirements.

A patient’s phone may hold only one record relevant to a study and may be available for training only during brief background execution windows. A suitable protocol should therefore require little local computation and allow clients to contribute without fitting local models or coordinating with other devices. Such participation could extend the remote data collection already used in decentralised clinical trials [6], and could support rare-disease studies in which small patient populations complicate recruitment [7]. The practical question is how to train from these individual contributions while accommodating interruptions and protecting the information exchanged during training.

Federated tree methods offer several ways to distribute this work. Recent surveys distinguish horizontal learning across different records from vertical learning across different features of overlapping records [8, 9]. In horizontal settings, SimFL and BOFRF rely on locally fitted models [10, 11], whereas FederBoost and FedTree support joint construction of boosted trees from distributed statistics [12, 13]. Aminifar et al. [14] jointly construct extremely randomized trees from protected class counts, using a different model and training statistic. Particularly close to our setting, Liu et al.’s original 2019 FedXGB addresses mobile crowdsensing through protected gradient aggregation and dropout handling, with evaluation on physical devices [15]. Maddock et al. [16] explicitly consider one record per participant and combine secure aggregation with differential privacy. Their framework protects released information as well as individual contributions during aggregation. Xu et al.’s PrivBoost combines client-generated noise with secure aggregation to train differentially private boosted trees. It assumes that institutional clients remain available throughout training and was evaluated on a workstation with emulated client–server communication [17]. Single-record learning and mobile federated boosting thus have established precedents. We investigate how a concrete aggregation protocol supports training contributions during brief, intermittent background execution on personal devices. Table 1 compares these approaches and related vertical methods by data setting, model, training organisation and privacy mechanism.

**Table 1:** Design choices in federated tree learning. GBDT = gradient-boosted decision trees; RF = random forest; ERT = extremely randomized trees; HE = homomorphic encryption; SA = secure aggregation; DP = differential privacy; LSH = locality-sensitive hashing; SS = secret sharing. Mechanisms have different adversary assumptions and permitted outputs; their inclusion does not imply equivalent privacy guarantees. FedTree’s mechanisms are configurable and setting-dependent. FedXGB refers to Liu et al.’s original 2019 arXiv version. BOFRF’s leaf-size restriction is a heuristic mitigation rather than a cryptographic or DP guarantee.

| **Method** | **Data partitioning** | **Model family** | **Training organisation** | **Privacy mechanism** |
|---|---|---|---|---|
| SecureBoost [18] | Vertical | GBDT | Joint training across feature holders | HE |
| Federated Forest [19] | Vertical | RF | Distributed construction with a trusted coordinator | Encryption |
| Xie et al. [20] | Vertical | GBDT | Joint computation across feature holders | SS; First-Layer-Mask |
| SimFL [10] | Horizontal | GBDT | Local trees informed by cross-party similarities | LSH; heuristic protection |
| BOFRF [11] | Horizontal | RF | Weighted combination of local forests | Local data retention; minimum leaf size in evaluation |
| Aminifar et al. [14] | Horizontal | ERT | Joint construction from class counts via a mediator | Masked SA |
| FederBoost [12] | Both | GBDT | Joint tree construction | Horizontal SA; vertical bucket randomisation |
| FedTree [13] | Both | GBDT | Joint histogram construction | SA, HE and DP |
| FedXGB [15] | Horizontal | GBDT | Joint training through edge servers | SS, masking and encryption |
| Maddock et al. [16] | Horizontal | GBDT | Joint training from protected aggregate statistics | SA and DP |
| PrivateBoost | Horizontal | GBDT | Joint training through fixed external shareholders | Threshold SS; optional path hiding |

We developed PrivateBoost around a fixed set of shareholders that aggregate contributions on behalf of personal devices (Fig. 1). Each device computes the statistics required for gradient boosting and distributes Shamir secret shares [21]; shareholders sum them, and an aggregator reconstructs the combined statistics used to build the trees. This arrangement avoids client-to-client coordination, unlike aggregation protocols that distribute masking secrets among participating clients [22, 23], while introducing assumptions about shareholder availability and collusion. Matching contributor sets across shareholders maintains consistent reconstruction when messages reach only some of them. An optional path-hiding mechanism conceals training-node membership. These protections concern the aggregation process: PrivateBoost does not provide the differential-privacy guarantee for released aggregates or models studied by Maddock et al. [16]. For a fixed model configuration and shareholder set, each client’s contribution size is independent of the total client population, with infrastructure parties handling the growing aggregation workload.

**Fig. 1:**
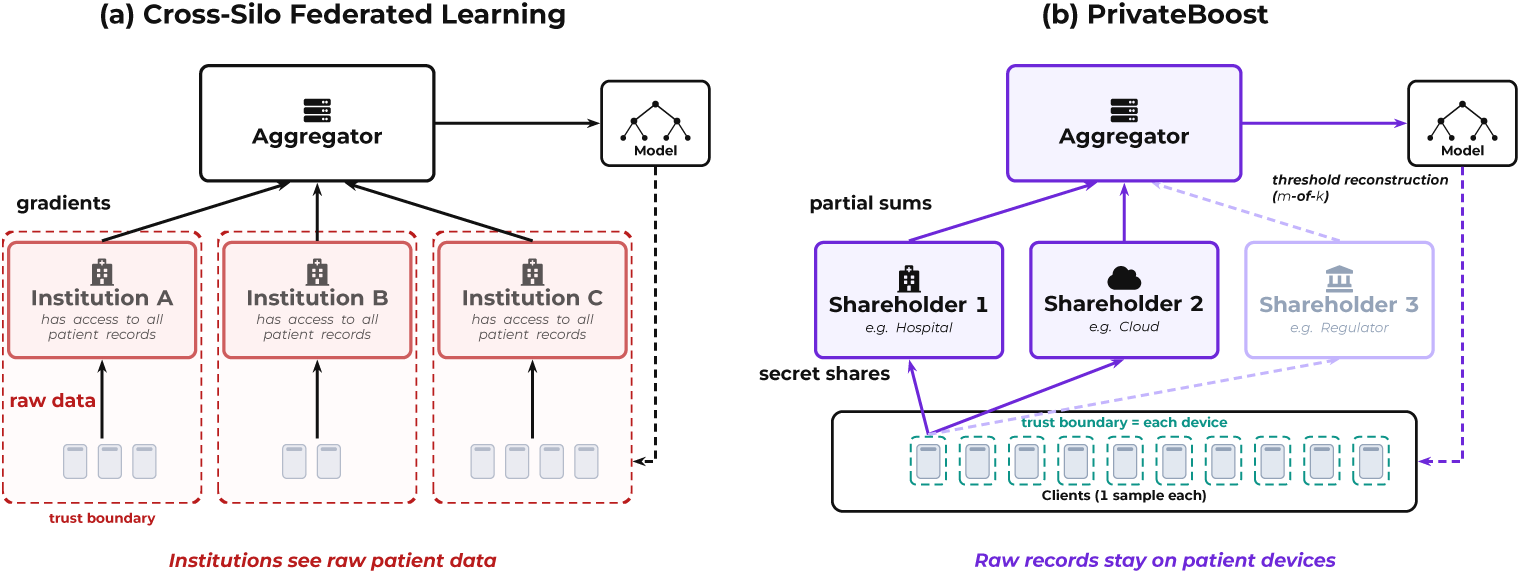
Institution-centric versus patient-level federated learning with PrivateBoost. (a) In the conventional cross-silo setting, patients provide data to institutions, which perform local training and share model updates. Each institution has access to the records within its cohort. (b) In PrivateBoost, individual patients can participate directly: each client holds a single record and distributes secret shares to a fixed set of shareholders. Shareholders aggregate shared statistics rather than raw records; protection depends on the stated collusion assumptions and does not exclude inference from released aggregates.

We evaluate this design through single-record learning simulations, communication measurements and a physical deployment. Simulations on four medical datasets assess predictive performance and resilience to specified dropout and message-loss patterns. Loopback measurements quantify per-client traffic under alternative sharing thresholds and path-hiding settings. Four consumer smartphones and tablets, each holding a batch of records, complete three training sessions unattended during operating-systemmanaged background execution. These evaluations establish single-record learning in simulation and mobile participation with low measured computation time. Together, they show how a concrete aggregation protocol can support contributions within the brief execution opportunities available on personal devices.

## Methods

### Problem statement

We consider a set of *c* clients 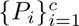, each holding a single labelled sample (**x***_i_, y_i_*) where **x***_i_* ∈ ℝ*^d^* is a feature vector and *y_i_* is the corresponding label. This one-sample-per-client setting arises naturally in medical applications where individual patients contribute their own health records without pooling raw records at an institution.

The problem of privacy-preserving federated XGBoost in the cross-device setting can be stated as follows:

#### Given

*c* clients, each holding a single sample (**x***_i_, y_i_*); a set of *n* shareholders; and a single aggregator.

#### Learn

An XGBoost ensemble of *T* trees from the distributed records 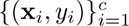 using securely aggregated statistics.

#### Security requirement

Protect individual contributions during aggregation, assuming that all parties follow the protocol, fewer than *m* shareholders collude, and no shareholder colludes with the aggregator. Here, *m* denotes the number of shares required for reconstruction. The protection permits the aggregate outputs and metadata described below; it does not prevent inference from that information. Specifically:

- The aggregator reconstructs aggregate gradient sums and feature statistics from summed shares, without receiving individual contribution shares or raw records.
- Any coalition of fewer than *m* out of *n* shareholders obtains no information about individual client contributions from their secret shares (information-theoretic guarantee via Shamir secret sharing).

#### Participation requirement

No client-to-client communication is required, accommodating intermittently available mobile devices.

The “Security analysis” section specifies the observations available to each party and the collusion assumptions underlying the analysis.

### Roles

We define three roles in the protocol:

**Definition 1** (Client) A client *P_i_* holds a single sample (**x***_i_, y_i_*). In each protocol round, the client computes its contribution (feature statistics or gradient–Hessian pairs), encodes these as Shamir secret shares, and distributes shares to all *n* shareholders. Clients receive broadcast information (bin edges, split decisions) from the aggregator, but do not send their records or individual training contributions to it directly. Clients do not communicate with each other.

**Definition 2** (Shareholder) A shareholder *S_j_* (*j* = 1*, …, n*) receives secret shares from clients and computes partial sums over received shares. Shareholders forward these partial sums to the aggregator upon request. Each shareholder receives one share of each client’s contribution, which is information-theoretically independent of the secret value.

**Definition 3** (Aggregator) The aggregator *A* coordinates the training process. It collects partial sums from at least *m* shareholders, reconstructs aggregate statistics via Lagrange interpolation, determines bin edges and split decisions, and broadcasts these to clients. The aggregator receives summed shares and round commitments, not individual contribution shares or raw records.

### Challenges

Our design addresses four requirements of single-record participation on intermittently available devices:

1. **Single-record contributions.** Each client holds exactly one sample, so local gradient computation produces a single (*g_i_, h_i_*) pair per round rather than a batch. There are no local statistics to aggregate before communication—every individual value must be protected.
2. **Histogram construction without raw data.** Histogram-based training uses common feature-bin boundaries across clients. In our setting, the aggregator has no access to pooled raw feature values, so we derive bin edges from securely aggregated statistics.
3. **Intermittent availability.** Cross-device clients (e.g., patient smartphones) may go offline at any time. The protocol must tolerate client dropout without corrupting aggregated results or requiring restarts, provided a sufficiently large common contributing cohort remains available.
4. **Path privacy.** As a tree is built, each client’s traversal path encodes information about its feature values (e.g., that a patient’s glucose exceeds a split threshold). Shareholders who observe which nodes a client contributes to can infer this path, creating a side channel not present in centralised training.

The protocol flow described below addresses challenges (1)–(3), the path-hiding extension addresses (4), and the security analysis describes the protections and their limits.

### Histogram-based split finding

XGBoost [24] builds decision trees by greedily selecting splits that maximise a gain function. The gain depends on sums of the firstand second-order derivatives of the loss evaluated at the current ensemble predictions. Without discretisation, the algorithm would need to evaluate every unique feature value as a candidate threshold. Histogrambased methods make this tractable by grouping continuous feature values into bins and evaluating candidate splits at their boundaries.

Each sample *i* contributes a gradient *g_i_* = *∂L/∂ŷ_i_* and Hessian 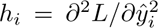 based on the current prediction *ŷ_i_* and true label *y_i_*. The gradient measures the local slope of the loss, and the Hessian measures its local curvature. For a candidate split that partitions samples into left (*L*) and right (*R*) sets, the gain is:

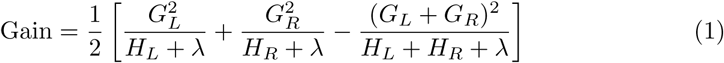

where *G_L_* = Σ*_i∈L_ g_i_* and *H_L_* = Σ*_i∈L_ h_i_*, with *G_R_* and *H_R_* defined analogously for the right child, and *λ* is a regularisation parameter. We use the split-gain formula of Chen and Guestrin [24], with the penalty for additional leaves set to zero. Candidate splits must satisfy a minimum total Hessian weight in each child node. Evaluating these splits requires only aggregate gradient and Hessian sums for each bin, which our protocol computes using secret sharing.

### System model

Fig. 2 summarises communication between clients, shareholders and the aggregator.

**Fig. 2:**
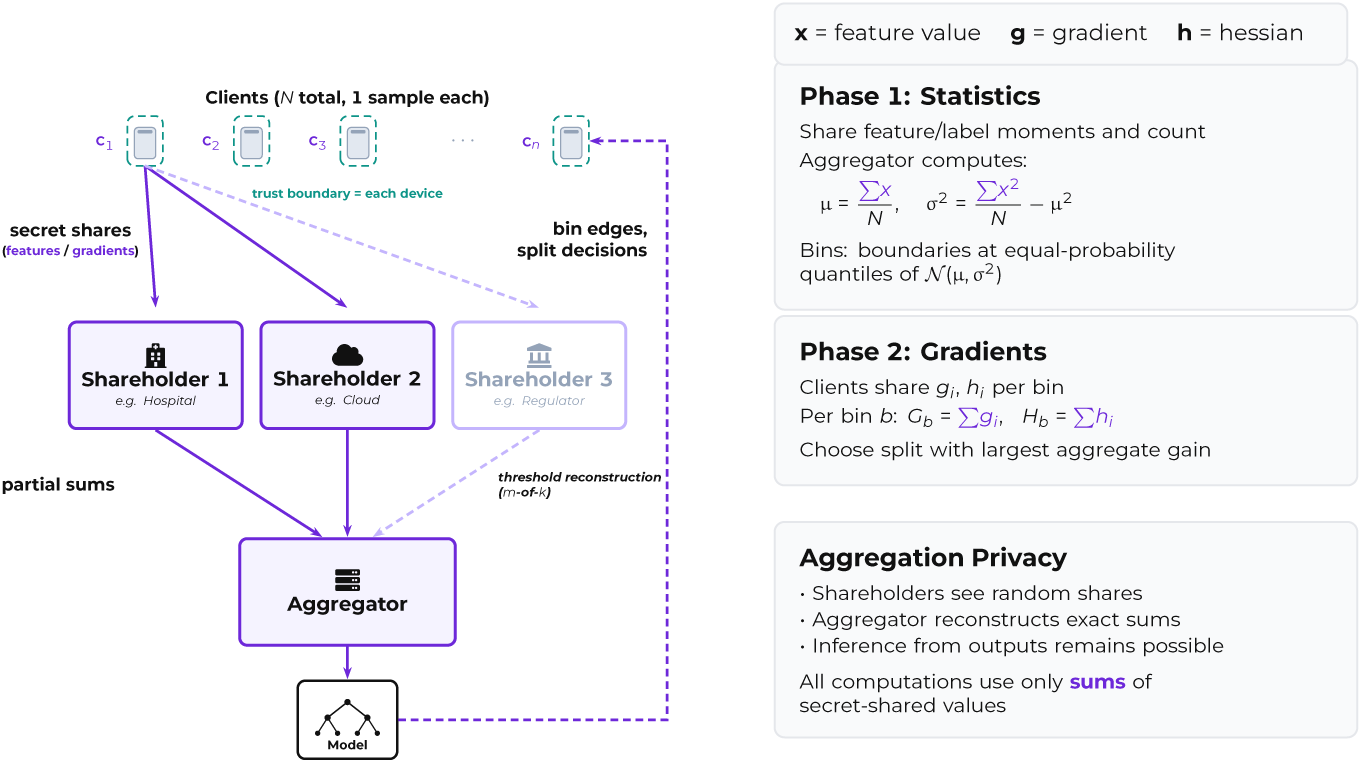
System architecture. Clients distribute Shamir shares to all shareholders. Shareholders sum received shares and forward partial sums to the aggregator, which reconstructs aggregate statistics (Σ*x*, Σ*x*^2^ for binning) and gradient sums (Σ*G*, Σ*H* for splits). The aggregator broadcasts bin configurations and split decisions back to clients.

### Building blocks

#### Shamir Secret Sharing

To share a secret value *s*, sample a random polynomial *f* (*x*) of degree at most *m* − 1 with *f* (0) = *s*. Distribute shares (*i, f* (*i*)) for *i* = 1*, …, n* to *n* shareholders. Any *m* shares can reconstruct *s* via Lagrange interpolation; fewer than *m* shares reveal nothing about *s*.

#### Finite Field Arithmetic

Shamir sharing requires arithmetic over a finite field F*_p_*. Following Catrina and Saxena [25], we represent real-valued gradients and statistics as fixed-point integers: we quantise a value *v* as ⌊*v* · 2*^k^*⌉ and encode it in the field as that integer modulo *p*, where *k* is the number of fractional bits. Negative values map to the upper half of the field (*p* − |⌊*v* · 2*^k^*⌉|). We choose *p* = 2^61^ − 1, a Mersenne prime that fits in a 64-bit integer and admits fast modular reduction via bit shifts. We use *k* = 24 fractional bits (≈7 decimal digits of resolution). Signed reconstruction recovers the integer sum exactly provided it lies in [−(*p* − 1)*/*2, (*p* − 1)*/*2] before reduction modulo *p*.

#### Additive Homomorphism

Adding shares of *a* and *b* held by the same shareholder produces a valid share of *a* + *b*. This allows shareholders to sum their received shares without learning individual values, while the aggregator reconstructs the aggregate sum.

#### Commitments

Each client generates a commitment *c* = *H*(round||client_id||nonce) using a fresh nonce per round. The aggregator selects the subset of *m* shareholders with the largest common set of commitments. These selected shareholders sum shares for that same set, provided it meets the minimum-client threshold *k*_min_. A client need not reach all *n* shareholders: its contribution is excluded if it is absent from any selected shareholder. If no eligible subset meets the threshold, the round cannot proceed to reconstruction. The aggregator sees only commitment hashes, not client identifiers.

### Protocol flow

The protocol has two phases: a one-time statistics phase to define histogram bins, followed by repeated gradient phases to build trees.

#### Phase 1: Statistics

Before training, the aggregator needs feature statistics to define histogram bin edges. Each client *i* creates Shamir shares of their feature values *x_i,f_* and squared values 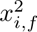 for each feature *f*, their label *y_i_* and squared label 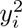, and a shared record count (one for a single-record client), along with a commitment. Clients send shares to shareholders. The aggregator selects a common contributing cohort as described above, requests its sums from the selected shareholders, reconstructs Σ*x* and Σ*x*^2^, and the record count, and derives per-feature mean *µ* and standard deviation *σ*. It also reconstructs the label sums. For logistic loss, the initial ensemble score is log(*y̅*/(1 − *y̅*)), where *y̅* is the cohort mean label clipped to [10*^−^*^7^, 1 − 10*^−^*^7^].

From these statistics the aggregator places *B* + 1 finite bin edges at equalprobability quantiles of N (*µ, σ*^2^):

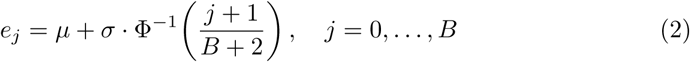

where Φ*^−^*^1^ is the inverse normal CDF, computed via rational approximation. These edges define *B* + 2 bins, including two outer bins extending to −∞ and +∞, respectively. Because the statistics phase already collects the necessary aggregates, Gaussian quantile binning adds no communication cost compared to a simpler equal-width scheme. The aggregator broadcasts the resulting bin edges to all clients.

#### Phase 2: Gradient Rounds

For each tree and each depth level, the aggregator needs gradient sums per (feature, bin) combination. Each client *i*:

1. Computes their gradient *g_i_* and Hessian *h_i_* from current ensemble prediction
2. Builds a gradient histogram spanning every bin of every feature: for each feature *f*, the bin containing *x_i,f_* holds (*g_i_, h_i_*) and every other bin holds (0, 0)
3. Creates Shamir shares of the entire histogram vector with a fresh commitment
4. Sends one share of the vector to each shareholder, tagged with the commitment

Each histogram entry is independently secret-shared, so a shareholder’s shares do not reveal whether that entry contains a real contribution or zero. Node tags may still reveal the client’s tree path, as discussed in “Path hiding”. For each active node, the selected shareholders sum the histogram vectors position-wise over the same contributing cohort. The aggregator reconstructs the gradient and Hessian sums for each feature bin, evaluates eligible splits at bin boundaries, and broadcasts the selected feature and threshold. Clients update their node assignments, and the process repeats for the next depth level.

##### Algorithm 1 PrivateBoost Training (with Path Hiding)

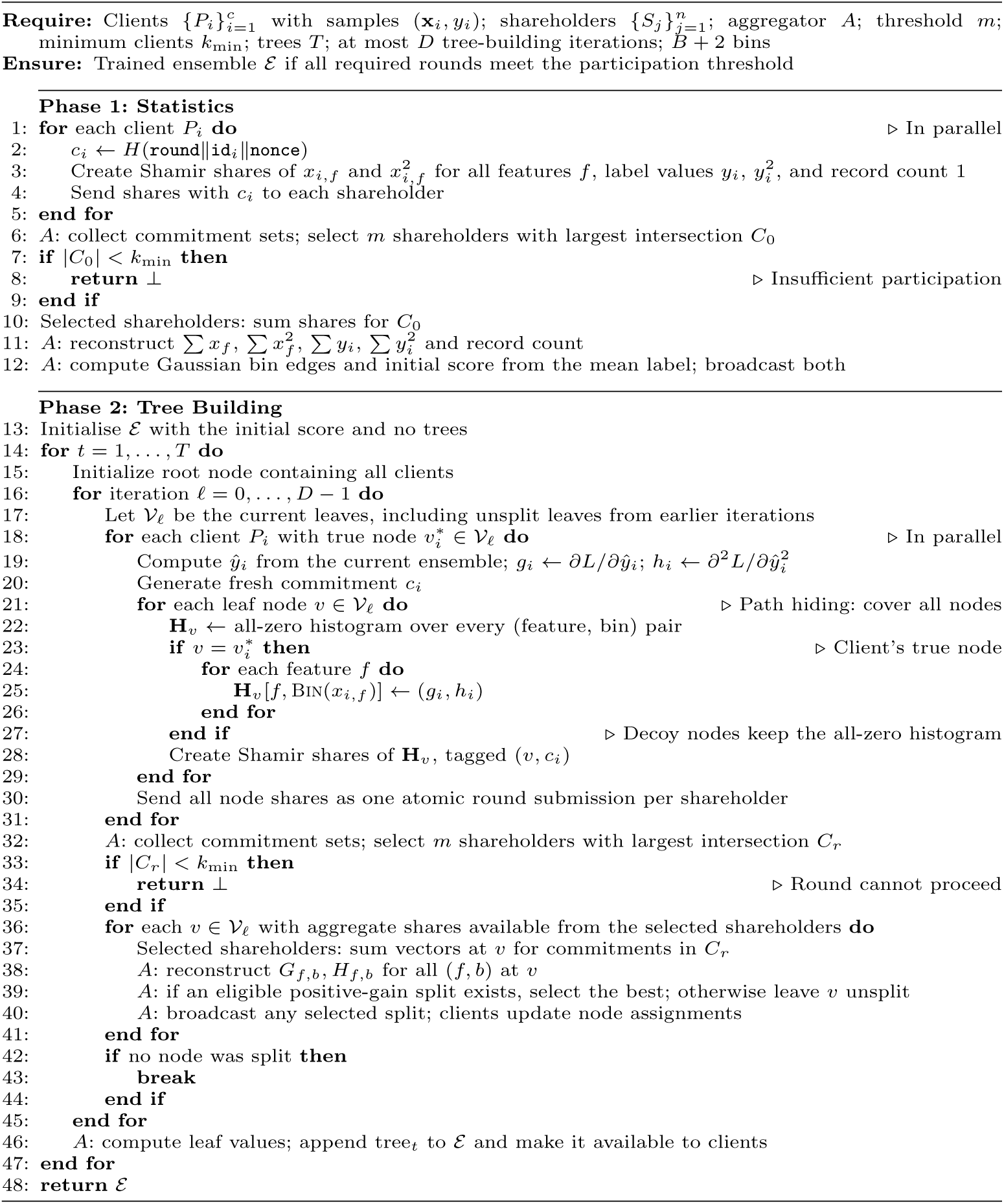

After the final tree-building iteration, or when no further splits are found, leaf values are computed from gradient sums, and the tree is added to the ensemble. The entire gradient phase repeats for each tree.

Algorithm 1 summarises the complete protocol.

### Path hiding

In the basic protocol, each client submits its histogram vector only for its current tree node. The shares themselves reveal nothing, but the node tag accompanying them does: shareholders learn which leaf the client occupies, and hence its tree path. For medical data, this can be sensitive—e.g., a shareholder learns that a patient’s glucose exceeds a diagnostic threshold.

Path hiding eliminates this leakage. Instead of submitting a vector for only its true node, each client submits vectors for *all* active leaf nodes in the current treebuilding iteration (Fig. 3). For the true node, the client shares its actual histogram vector. For all other nodes, it shares an all-zero vector. Since Shamir shares of zero are indistinguishable from shares of any other value, a coalition of fewer than *m* shareholders cannot distinguish the client’s true node from the shared vectors alone.

**Fig. 3:**
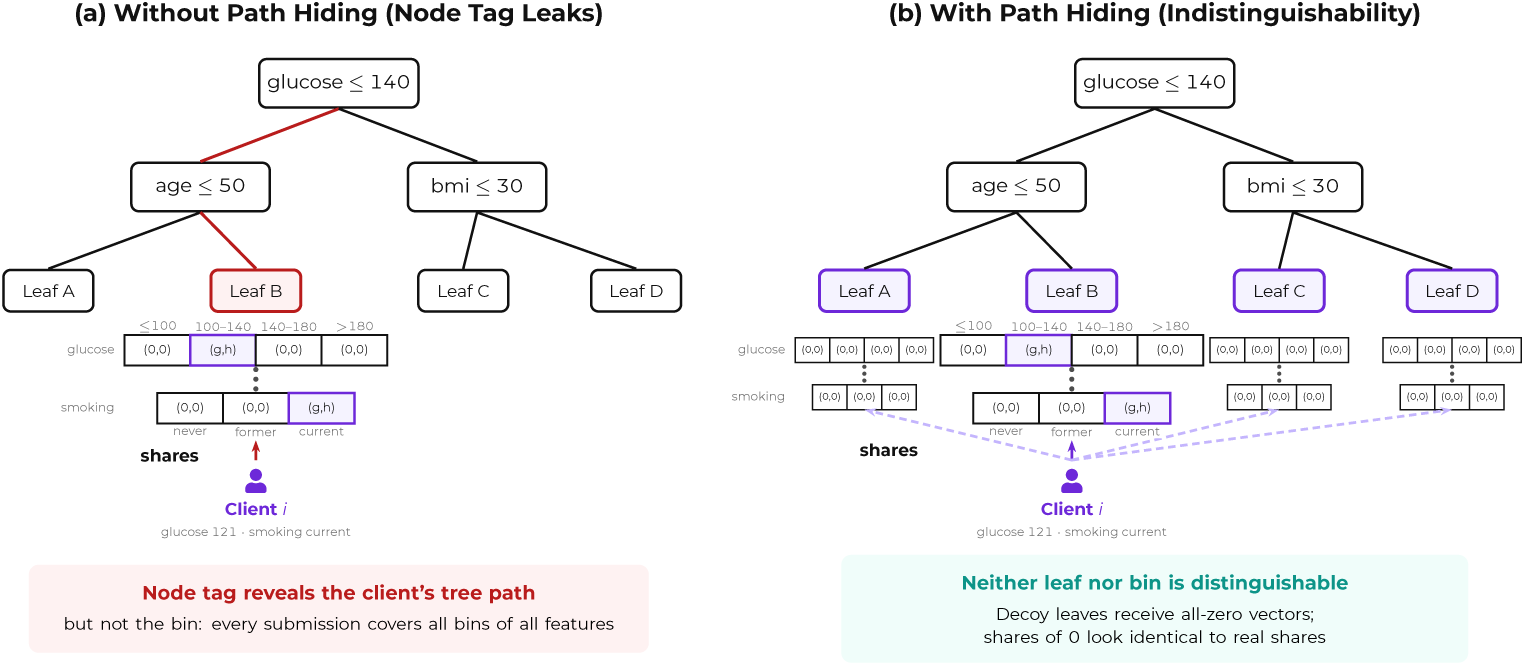
Path hiding. Each submission is a dense bin vector spanning every bin of every feature, shown for a numerical feature (glucose) and a categorical one (smoking status); the gradient–Hessian pair occupies the true bin of each feature, and all other bins hold (0, 0). (a) Without path hiding, the client submits the vector only for its true leaf node, whose node tag reveals its tree path. (b) With path hiding, the client submits a vector for every active leaf node; decoy nodes receive shares of all-zero vectors.

For the same participating records and training decisions, adding secret-shared zero contributions for unoccupied nodes leaves the reconstructed aggregates unchanged. Path hiding therefore preserves the resulting model under these conditions. The cost is communication overhead: in tree-building iteration *ℓ*, each client submits vectors for |V*_ℓ_*| ≤ 2*^ℓ^* active nodes instead of one (quantified in “Communication cost” in Results). This mechanism is captured by the loop over active leaf nodes in Algorithm 1. Disabling path hiding reduces to submitting shares only for *v^∗^*.

### Security analysis

PrivateBoost protects individual contributions during aggregation using threshold secret sharing. We analyse an honest-but-curious setting in which parties follow the protocol but may attempt to infer private information from their observations. We consider the aggregator, a coalition of fewer than *m* shareholders, and a coalition of clients separately. The analysis does not cover coalitions combining these roles or parties that deviate from the prescribed computation. This analysis identifies information exposed by the protocol and potential attack mechanisms; it does not empirically evaluate attack effectiveness.

Each client distributes its encoded contribution among *n* shareholders using Shamir secret sharing. With independently and uniformly sampled polynomial coefficients, any collection of fewer than *m* shares has a joint distribution independent of the shared value [21]. This property applies separately to every entry of the feature-statistics and gradient–Hessian vectors. Shareholders can therefore add their shares without recovering individual contributions, while the aggregator reconstructs sums from the responses of *m* shareholders. The information-theoretic property concerns the mathematical sharing scheme; its implementation uses a pseudorandom generator to sample coefficients.

The reconstruction threshold determines the tolerated shareholder collusion. Our default 2-of-3 configuration protects individual shares against one curious shareholder, whereas 3-of-5 protects against coalitions of up to two. A coalition of *m* shareholders can reconstruct an individual contribution. Aggregation requires the selected shareholders to hold submissions from a common cohort of at least *k*_min_ clients; subject to this condition, up to *n* − *m* shareholders may be unavailable. Increasing *n* at fixed *m* provides more choices of an available shareholder subset but does not raise the collusion threshold. Increasing *m* strengthens collusion resistance while requiring more shareholder responses. Each client sends shares to all *n* shareholders, so increasing their number also increases communication. Table 6 reports the measured costs of both configurations.

Protection of the numerical shares does not conceal all information associated with a submission. Each shareholder observes persistent client pseudonyms and can link participation across rounds. In the base protocol, node tags also reveal the client’s current tree node and hence its path through the tree. Individual feature-bin assignments are not transmitted as metadata: each histogram contains shares for every bin, including shares of zero for unoccupied positions. Under the shareholder threshold assumption, these shares do not distinguish occupied from empty bins beyond information available from the visible metadata. Path hiding extends this construction to every active node, so node tags no longer identify the client’s actual node. It does not conceal participation, persistent pseudonyms or transport metadata.

The aggregator receives aggregate outputs rather than individual contribution shares. During the statistics phase, these outputs comprise feature and label sums, their squared-value sums, and the contributing record count. They disclose population summaries, including feature means and the prevalence of the positive label in the cohort. These are intended outputs, but may still be sensitive. During training, the aggregator receives gradient and Hessian sums for each active node and feature bin. It also observes round commitments, their delivery across shareholders and the selected shareholder set. Fresh nonces make commitments vary across rounds, but do not hide the delivery pattern within a round. Clients receive the bin edges, initial ensemble score, split decisions and updated ensemble, including leaf values.

Small node or bin populations can expose individual information within these aggregates. The minimum-client threshold applies to the common contributing cohort, not to occupancy within each node or bin. Although bin counts are not separately released, they can sometimes be recovered from the Hessian sums. Under binary logistic loss with a common predicted probability of 0.5, each record contributes a Hessian of 0.25, making the bin count four times its Hessian sum. If that count is one, the gradient *g_i_* = 0.5 − *y_i_* also reveals the record’s binary label: +0.5 corresponds to *y_i_* = 0 and −0.5 to *y_i_* = 1. Associating the recovered label with a particular person requires additional information. Contributions disclosed by colluding clients could likewise be subtracted from an aggregate, potentially isolating another client’s contribution; such mixed coalitions are outside the analysis.

Repeated releases create further opportunities for inference. If two aggregates contain identical contributions from the same records except for one, their difference reveals that record’s contribution even when both cohorts exceed *k*_min_. Changing gradients and unknown cohort overlap complicate this subtraction, and attributing the result to an individual requires further information. Lam et al. demonstrated disaggregation of federated updates using repeated aggregates and participation summary analytics, assuming an adversarial server that can hold the model fixed across rounds [26]. This motivates considering releases jointly. Controls on repeated aggregation requests and cohort selection could limit opportunities for deliberate isolation, but a minimum cohort size alone does not protect against inference across releases.

Secure aggregation therefore limits direct access to individual gradients without establishing protection against reconstruction from released statistics or membership inference from trained models. Gradient reconstruction and membership inference have been demonstrated in neural-network settings [27, 28]. Differential privacy offers complementary protection by limiting the influence of an individual record on released information and has been applied to federated boosted trees [16]. An extension would need bounded contributions, calibrated noise and privacy accounting across the statistics phase and subsequent training releases. Protection against the aggregator would require perturbation before it observes the sums. PrivateBoost does not currently implement differential privacy.

Malicious behaviour introduces risks beyond this semi-honest analysis. A coordinator could attempt to isolate contributions by manipulating cohort selection or providing different training contexts to different clients. Pasquini et al. demonstrated model-inconsistency attacks against secure aggregation in federated neural-network training [29]. Verifiable agreement on the training context and enforceable aggregation rules would be relevant safeguards. If enrolment permits an attacker to register multiple client identities, those identities could dominate a cohort, distort training or facilitate subtraction using their known contributions. The client-count threshold does not establish that participants are independent people. Enrolment controls would address this concern, while validation of contribution bounds and histogram structure would address malformed submissions. These safeguards would require separate design and evaluation; the current analysis establishes neither resistance to malicious coordination nor protection against multiple attacker-controlled identities. Contribution validation and attestation are discussed further in the Discussion.

### Communication and computation cost

We give bounds for single-record clients. Let *c* denote the number of clients, *n* the number of shareholders, *m* the reconstruction threshold, *F* the number of features, *T* the number of trees, and *d* the maximum number of tree-building iterations. Let V*_t,ℓ_* denote the current leaves considered in iteration *ℓ* of tree *t*, taking this set to be empty for iterations omitted by early stopping. There are *B* + 2 logical bins, including overflow bins. The current implementation allocates *B*_slots_ = *B* + 3 histogram entries per feature, using the boundary-array length; the final entry is unused for finite feature values. Write 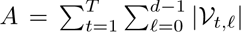 for the total number of active-node processing steps, with *A* ≤ *T* (2*^d^* − 1).

In the base protocol, each single-record client submits a dense gradient–Hessian histogram for at most one node per tree-building iteration. Client-to-shareholder histogram traffic is therefore *O*(*TdFB*_slots_*cn*) field elements. With path hiding, each client submits for every active node, giving an upper bound of *O*(*AFB*_slots_*cn*). The one-time statistics phase adds *O*(*Fcn*) field elements. For each active node with sufficient aggregate shares, the selected *m* shareholders return summed histogram shares, giving at most *O*(*AFB*_slots_*m*) field elements over training. Split decisions are made separately for each node with an eligible split.

Sharing each secret requires evaluating a degree-(*m* − 1) polynomial at *n* points. Per-client sharing work is *O*(*TdFB*_slots_*nm*) in the base protocol and at most *O*(*AFB*_slots_*nm*) with path hiding. Shareholders add incoming shares position-wise. The implementation computes Lagrange coefficients once per reconstructed vector, costing *O*(*m*^2^) field operations, and then uses them for its *O*(*FB*_slots_) entries. Aggregate reconstruction therefore costs *O*(*A*(*m*^2^ + *FB*_slots_*m*)), with a further *O*(*AFB*_slots_) for split finding.

These expressions describe histogram traffic and the associated sharing, reconstruction and split-finding work. They exclude commitment exchange, shareholder selection, HTTP framing, polling and delivery of the model and round context; they do not represent total client computation. The current selection routine enumerates the 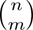 shareholder subsets and compares their commitment intersections; its cost matters when increasing the shareholder population. For fixed *m*, client sharing costs grow linearly with *n*, while aggregate reconstruction uses only *m* shareholders. More shareholders provide additional choices of an available subset, provided that subset has a sufficiently large common contributing cohort. Raising *m* improves collusion resistance but increases sharing and reconstruction costs. For fixed model and shareholder parameters, each single-record client’s histogram payload and sharing work are bounded independently of the total client population. “Communication cost” and “Real-world deployment” report measured traffic and device-side costs.

### Experimental setup

We evaluate whether privacy-preserving aggregation from single-record clients can match the predictive quality of centralised training across four structured medical classification tasks of increasing scale. Three small datasets long distributed by the UCI Machine Learning Repository represent common diagnostic scenarios: Heart Disease [30, 31] (Cleveland subset, 297 of 303 records retained after excluding those with missing values, 13 features), Breast Cancer [32, 33] (569 samples, 30 features, analysed from the copy bundled with scikit-learn), and Pima Diabetes [34, 35] (768 samples, 8 features). To evaluate predictive performance with a larger simulated population of single-record clients, we additionally use the CDC Diabetes Health Indicators dataset as distributed by UCI [36]. From the 253,680 records of that release we draw a classbalanced subset of 70,692 samples, comprising all 35,346 records labelled positive in the binary target together with an equal-sized random sample of negative cases, over 21 health indicator features predicting the combined prediabetes-or-diabetes binary target. In the predictive-performance simulations, each client holds exactly one training record, the regime in which a patient contributes a single diagnostic measurement from a personal device. We report results over 5 stratified random 80/20 train/test splits (mean ± std).

The evaluations use three distinct client configurations. Predictive-performance simulations assign one training record to each client. Loopback communication measurements use one single-record client and one batch client. The physical pilot uses four smartphones and tablets, each holding a batch, with the server roles co-located. The minimum-client threshold is separate from the Shamir reconstruction threshold: the simulation executable defaults to *k*_min_ = 10; the loopback session requires two clients, with shareholder release floors set to one for functional measurement; and the pilot configuration sets both the session minimum and shareholder release floor to three devices. These thresholds apply to the common contributing cohort, not to the number of nonzero contributions in each node or bin.

We use a 2-of-3 Shamir threshold, 10 bins per feature, max depth 3, 15 trees, learning rate 0.15, regularisation *λ* = 2.0, and logistic loss. Bin edges are placed at Gaussian quantile boundaries as described in “Protocol flow”. We compare against two XGBoost baselines: (1) XGBoost with matched tree hyperparameters (depth, trees, learning rate, *λ*, and 10 bins), and (2) XGBoost with default settings.

## Results

### Model quality

PrivateBoost achieved predictive performance comparable to matched centralised XGBoost across the four evaluated datasets (Fig. 4). The learning curves overlap or closely track the matched configuration, typically reaching their final AUC-ROC within 8–10 trees.

**Fig. 4:**
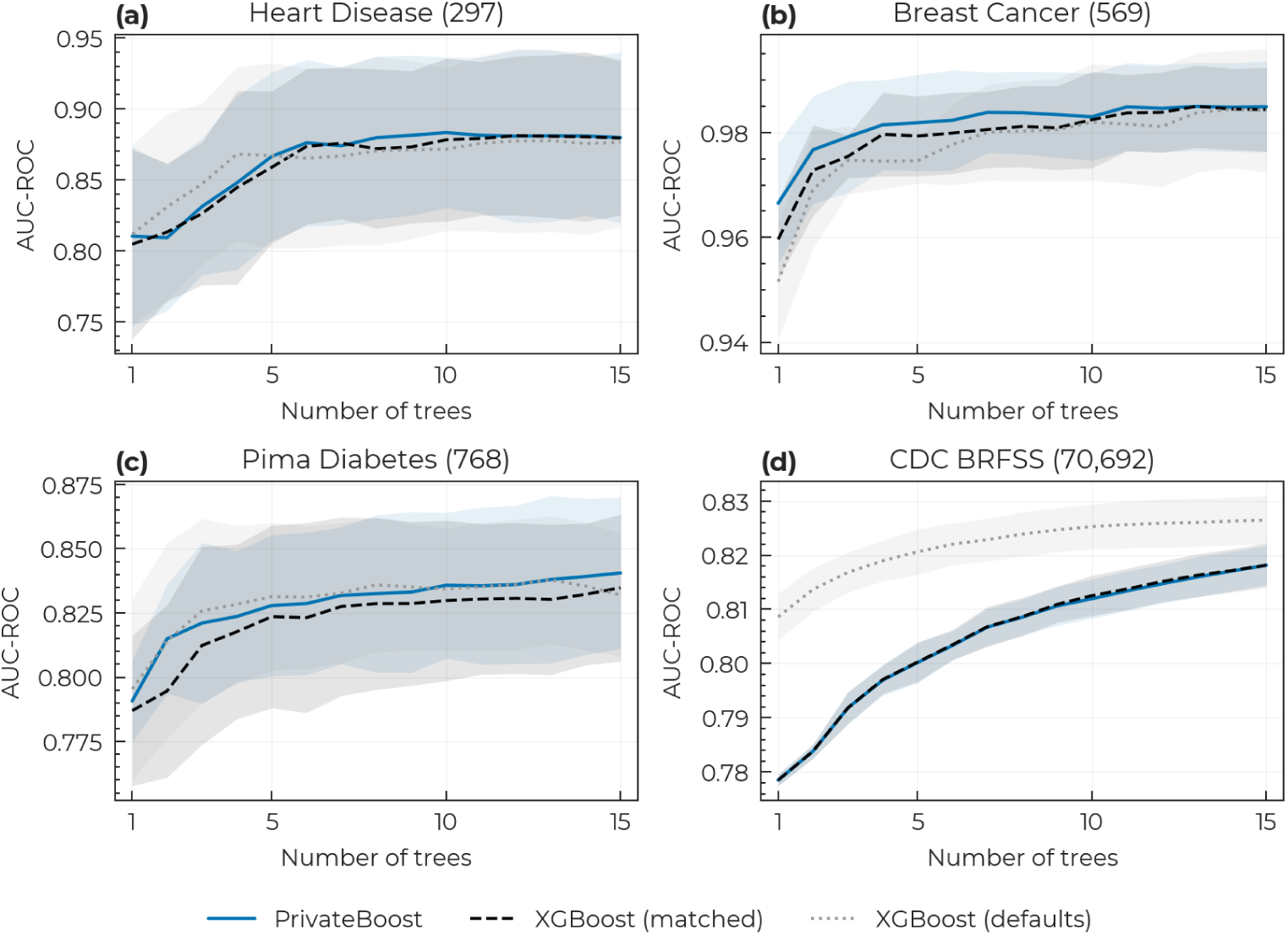
Model quality versus ensemble size on four medical datasets. AUCROC against the number of boosting trees for PrivateBoost, XGBoost with matched hyperparameters, and XGBoost with defaults, on (a) Heart Disease, (b) Breast Cancer, (c) Pima Diabetes and (d) CDC BRFSS; the number in parentheses in each panel title is the total number of records before the train/test split. Lines show the mean, shaded bands ±1 s.d. (*n* = 5 independent stratified splits).

Table 2 reports final classification metrics. PrivateBoost and matched XGBoost have the same mean accuracy on Heart Disease and CDC BRFSS (81.7% and 74.2%, respectively). Their mean AUC-ROC values are 0.880 and 0.879 on Heart Disease, respectively, and both are 0.818 on CDC BRFSS. Mean AUC-ROC is also similar on Breast Cancer (0.985 versus 0.984) and Pima Diabetes (0.841 versus 0.835). The default XGBoost configuration achieves a higher mean AUC-ROC of 0.826 on CDC BRFSS.

**Table 2:**
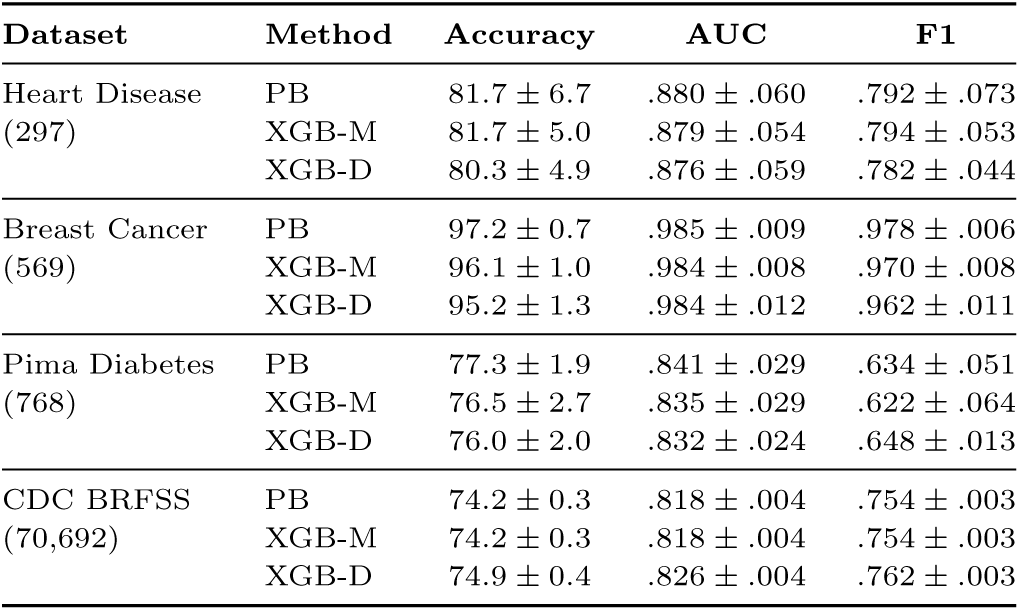
Classification metrics on the four medical datasets. Mean ± s.d. over *n* = 5 independent stratified splits; the number in parentheses under each dataset name is the total number of records before the train/test split. PB = PrivateBoost, XGB-M = XGBoost matched hyperparameters, XGB-D = XGBoost defaults.

| **Dataset** | **Method** | **Accuracy** | **AUC** | **F1** |
|---|---|---|---|---|
| Heart Disease<br>(297) | PB | 81.7 ± 6.7 | .880 ± .060 | .792 ± .073 |
|  | XGB-M | 81.7 ± 5.0 | .879 ± .054 | .794 ± .053 |
|  | XGB-D | 80.3 ± 4.9 | .876 ± .059 | .782 ± .044 |
| Breast Cancer<br>(569) | PB | 97.2 ± 0.7 | .985 ± .009 | .978 ± .006 |
|  | XGB-M | 96.1 ± 1.0 | .984 ± .008 | .970 ± .008 |
|  | XGB-D | 95.2 ± 1.3 | .984 ± .012 | .962 ± .011 |
| Pima Diabetes<br>(768) | PB | 77.3 ± 1.9 | .841 ± .029 | .634 ± .051 |
|  | XGB-M | 76.5 ± 2.7 | .835 ± .029 | .622 ± .064 |
|  | XGB-D | 76.0 ± 2.0 | .832 ± .024 | .648 ± .013 |
| CDC BRFSS<br>(70,692) | PB | 74.2 ± 0.3 | .818 ± .004 | .754 ± .003 |
|  | XGB-M | 74.2 ± 0.3 | .818 ± .004 | .754 ± .003 |
|  | XGB-D | 74.9 ± 0.4 | .826 ± .004 | .762 ± .003 |

### Binning analysis

In the evaluated implementation, PrivateBoost derives histogram boundaries from aggregate feature means and variances. These boundaries determine the candidate split thresholds available during training. We compare two strategies: Gaussian quantile binning, which places edges at equal-probability quantiles of N (*µ, σ*^2^), and equal-width binning, which spaces edges at equal intervals across the same range.

Table 3 quantifies bin uniformity using the coefficient of variation (CV) of bin occupancy, where CV = 0 corresponds to perfectly uniform bins. Gaussian quantile binning produces more uniform occupancy on continuous features (mean CV 0.59 vs. 0.66), while occupancy CVs are identical for discrete features. Table 3 also reports a paired feature-level Wilcoxon test; correlations between features within a dataset limit its inferential interpretation.

**Table 3:** Bin occupancy uniformity by feature type. Mean CV = *σ/µ* of bin counts, with lower values indicating more uniform occupancy, pooled across all four datasets; feature counts are given in parentheses. The *p*-value is from a two-sided Wilcoxon signed-rank test (paired by feature). A dash indicates that the test is undefined: the two methods give identical CVs for every discrete feature, leaving no non-zero paired differences to rank. Those tied pairs are dropped from the pooled test, so the value reported for all features is that of the continuous ones.

| **Features** | **Gaussian** | **Equal-width** | *p* |
|---|---|---|---|
| Cont. (45) | 0.59 | 0.66 | 9.6×10<sup>−8</sup> |
| Disc. (27) | 2.24 | 2.24 | — |
| All (72) | 1.20 | 1.25 | 9.6×10<sup>−8</sup> |

We assess split quality through gain retention: the ratio of the chosen binned split gain to the best gain obtained by evaluating all distinct-value thresholds for that same feature. Table 4 summarises retention within each run before comparing the methods across matched training splits. For continuous features, Gaussian binning has higher mean retention on Heart Disease, whereas equal-width binning has higher means on the other three datasets. Discrete-feature mean retention ranges from 0.976 to 1.000. Thus, more uniform bin occupancy does not consistently yield higher gain retention in these runs. These descriptive comparisons concern the nodes and features selected by each method, which can differ as training progresses; they do not establish equivalence between the methods.

**Table 4:** Gain retention by dataset and feature type. Retention is the ratio of the chosen binned split gain to the best exact-threshold gain for the same feature at that node. Node-level ratios are averaged within each training run and feature-type group, then summarised as mean ± sample s.d. across five matched train/test splits (15 trees, maximum depth 3, bin parameter *B* = 10). Each run has equal weight. Δ is the paired run-level difference (Gaussian minus equal-width), also reported as mean ± s.d. Continuous features have *>*20 unique values; discrete features have ≤20. Breast Cancer has no discrete features. The standard deviations describe variation across overlapping splits, not confidence intervals; no hypothesis test is applied.

| **Dataset** | **Features** | **Gaussian** | **Equal-width** | **Δ** |
|---|---|---|---|---|
| Heart Disease | Continuous | 0.903 ± 0.033 | 0.887 ± 0.045 | 0.016 ± 0.043 |
| Heart Disease | Discrete | 1.000 ± 0.000 | 1.000 ± 0.000 | 0.000 ± 0.000 |
| Heart Disease | All | 0.955 ± 0.012 | 0.950 ± 0.020 | 0.004 ± 0.018 |
| Breast Cancer | Continuous | 0.869 ± 0.025 | 0.884 ± 0.032 | −0.015 ± 0.042 |
| Breast Cancer | All | 0.869 ± 0.025 | 0.884 ± 0.032 | −0.015 ± 0.042 |
| Pima Diabetes | Continuous | 0.816 ± 0.020 | 0.829 ± 0.033 | −0.014 ± 0.040 |
| Pima Diabetes | Discrete | 0.976 ± 0.036 | 1.000 ± 0.000 | −0.024 ± 0.036 |
| Pima Diabetes | All | 0.835 ± 0.014 | 0.850 ± 0.028 | −0.015 ± 0.036 |
| CDC BRFSS | Continuous | 0.967 ± 0.012 | 0.972 ± 0.004 | −0.004 ± 0.011 |
| CDC BRFSS | Discrete | 1.000 ± 0.001 | 0.999 ± 0.002 | 0.001 ± 0.001 |
| CDC BRFSS | All | 0.988 ± 0.004 | 0.990 ± 0.002 | −0.002 ± 0.005 |

Fig. 5 illustrates the mechanisms on two continuous Pima Diabetes features. On near-symmetric Glucose (*γ*_1_ = 0.17), Gaussian quantile binning distributes bin edges more evenly across the data-dense region. On the heavily skewed Insulin feature (*γ*_1_ = 2.27), both methods concentrate approximately 49% of samples in a single bin due to a zero-inflated spike, limiting the resolution of either approach.

**Fig. 5:**
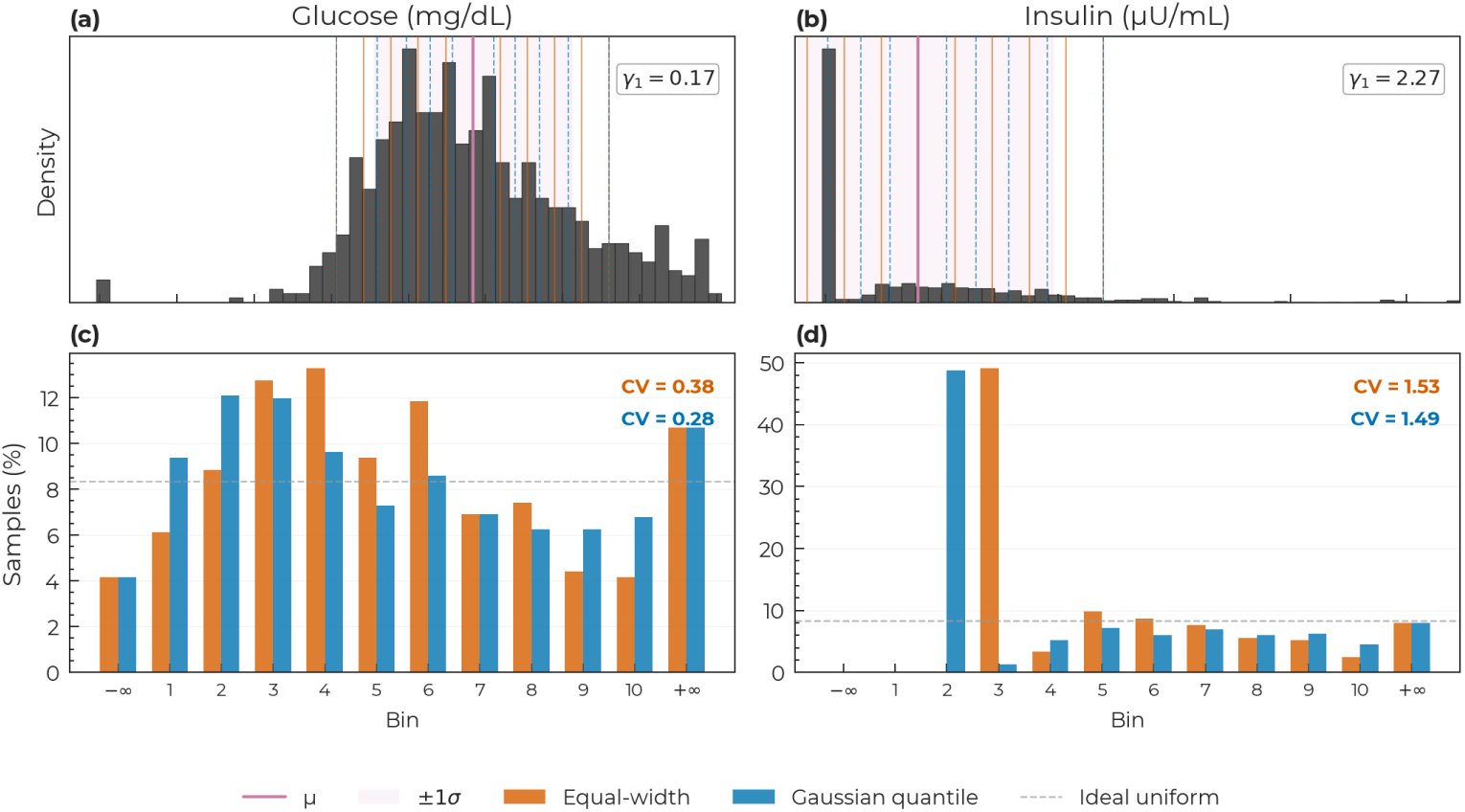
Bin occupancy under equal-width and Gaussian quantile binning. Two Pima Diabetes features of contrasting skewness (*γ*_1_) are shown. (a, b) Distributions of glucose (a) and insulin (b), with equal-width (amber, solid) and Gaussian quantile (blue, dashed) bin edges overlaid; the pink line marks the feature mean *µ* and the shaded band ±1*σ*. (c, d) Per-bin sample fractions under both methods for glucose (c) and insulin (d) (grouped bars), with the coefficient of variation (CV) of bin occupancy annotated for each; the dashed grey line is uniform occupancy. Bins −∞ and +∞ are the overflow bins outside the binned range.

Bin boundaries could also be specified using prior knowledge or an independent reference dataset. For example, established clinical thresholds for a blood test could define candidate split points without estimating them from participating clients’ records. The protocol would then aggregate gradients and Hessians within these predefined bins and select splits using the same gain criterion. The usefulness of such boundaries would depend on their relevance to the target population and prediction task.

### Client dropout resilience

Intermittent client availability can affect both bin estimation and tree construction. We evaluate three failure modes separately: dropout during gradient rounds, dropout during the one-time statistics phase, and loss of individual client-to-shareholder submissions. Aggregation can proceed when *m* shareholders hold contributions from a common cohort of at least *k*_min_ clients. The selected shareholders sum shares from that same cohort, ensuring consistent reconstruction despite missing submissions. The Shamir threshold *m* determines how many shareholder responses are required; *k*_min_ determines the minimum contributing cohort size and does not ensure minimum occupancy in individual nodes or bins. The following experiments assess predictive performance under randomly sampled failures.

#### Gradient-round dropout

We randomly exclude a fraction of clients each gradient round, with independent sampling per round. Mean AUC-ROC remains close to or above the no-dropout baseline on Heart Disease, Breast Cancer and CDC BRFSS at 80–90% dropout (Fig. 6). Pima Diabetes shows a modest reduction, from 0.841 without dropout to 0.828 and 0.831 at 80% and 90% dropout, respectively. Variation across training splits does not increase consistently across datasets. Because clients are resampled each round, records excluded from one round can contribute later; these results do not represent permanent loss of the same clients or dropout correlated with their records.

**Fig. 6:**
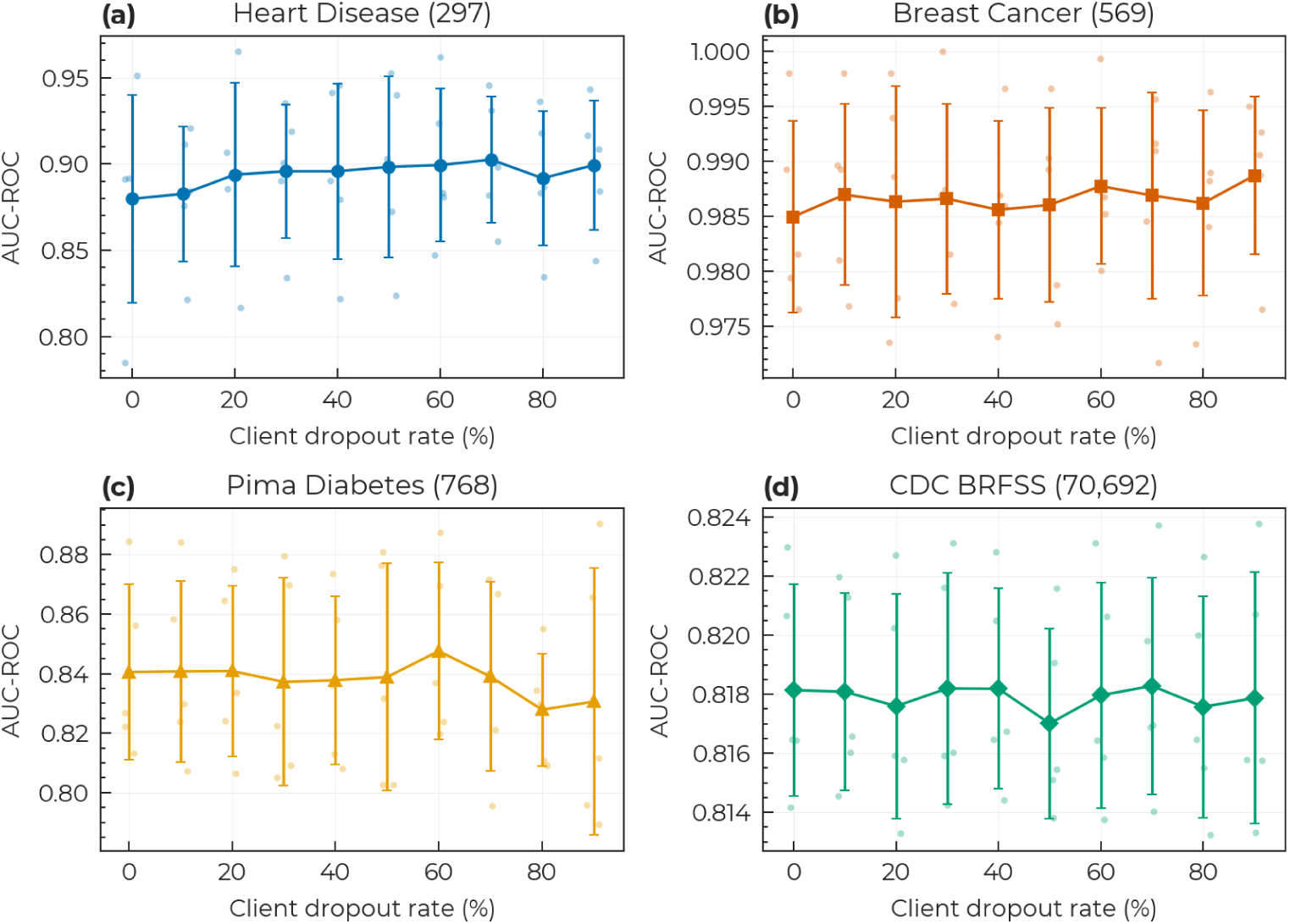
Model quality under client dropout. AUC-ROC against the fraction of clients excluded from each gradient round, on (a) Heart Disease, (b) Breast Cancer, (c) Pima Diabetes and (d) CDC BRFSS (2-of-3 threshold, 15 trees, depth 3). Data are mean ± s.d.; translucent points show individual values (*n* = 5 independent stratified splits).

#### Statistics-phase dropout

Table 5 reports AUC-ROC when bin edges are derived from a random subset of clients. Mean AUC-ROC remained close to the no-failure baseline at statistics-phase dropout rates of up to 90%. These results suggest that the aggregate mean and variance estimates were sufficient for binning in the evaluated settings.

**Table 5:** Model quality under statistics-phase dropout and per-shareholder message loss. AUC-ROC, mean ± s.d. over *n* = 5 independent stratified splits. Message loss is applied independently per (client, shareholder, round) submission. 2-of-3 threshold, 15 trees, max depth 3.

| **Dataset** | **No failures** | **Stats dropout** |  | **Message loss** |  |
|---|---|---|---|---|---|
|  |  | **50%** | **90%** | *q*=0.3 | *q*=0.5 |
| Heart Disease | .880 ± .060 | .889 ± .055 | .882 ± .056 | .892 ± .041 | .881 ± .032 |
| Breast Cancer | .985 ± .009 | .987 ± .007 | .986 ± .008 | .989 ± .005 | .984 ± .008 |
| Pima Diabetes | .841 ± .029 | .842 ± .028 | .836 ± .032 | .841 ± .025 | .832 ± .029 |
| CDC BRFSS | .818 ± .004 | .818 ± .003 | .818 ± .004 | .818 ± .004 | .818 ± .004 |

#### Per-shareholder message loss

Instead of removing whole clients, each clientto-shareholder submission is lost independently with probability *q* in every round. This exercises the consistency mechanism described in “Building blocks”: the aggregator selects the *m* shareholders whose received commitment sets have the largest intersection, and a client whose submission missed any of the selected shareholders is excluded from that round’s aggregate. Under this message-loss model, matching the contributing cohort prevents partial delivery from producing inconsistent aggregate shares; a round can proceed only if the common cohort meets the participation threshold. For any fixed pair of shareholders, *q* = 0.5 gives a contribution a (1−*q*)^2^ = 0.25 probability of reaching both; selecting the pair with the largest common cohort can retain more clients. In the evaluated 2-of-3 configuration, mean AUC-ROC decreases by at most 0.009 at *q* = 0.5 (Table 5), consistent with the gradient-round dropout results above.

### Communication cost

We measured per-client communication using the complete protocol with clients, shareholders and aggregator running on the same computer and communicating through the local loopback interface. Measurements cover all four datasets, both Shamir thresholds and both path-hiding settings. One client holds a single record, while a second holds the remaining dataset records as a batch. In each gradient round, clients send each shareholder a batch containing a commitment, routing information and histogram shares for their current node or, with path hiding, all active nodes. We record the single-record client’s total bytes sent and received at the TCP-stream interface, including TLS handshakes, HTTP/2 framing, submissions and round-context polling. These measurements provide a controlled comparison of per-client traffic across protocol configurations. The physical-device pilot below evaluates participation and submission costs during mobile background execution.

Without path hiding, total per-client traffic under the 2-of-3 configuration ranges from 586 KiB for Pima Diabetes to 1,368 KiB for Breast Cancer (Table 6). Traffic includes both submitted shares and downloaded round context: for CDC BRFSS, downloads account for approximately 222 KiB of the 1,056 KiB total. Moving to 3-of5 increases traffic by approximately 1.4–1.5 times, as clients distribute shares to more shareholders. These totals describe a single training run under the measured polling schedule.

**Table 6:**
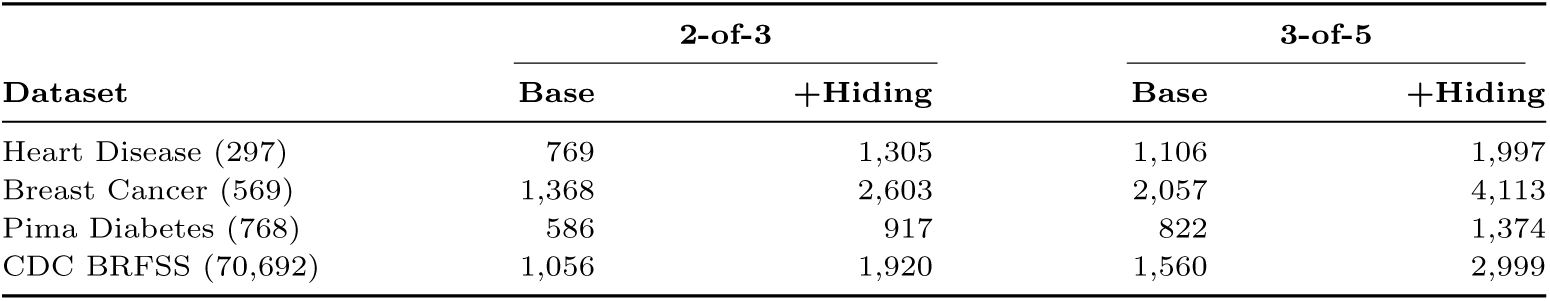
Per-client communication cost under different Shamir thresholds and path-hiding settings. Kibibytes (KiB) sent and received by one client’s device over a complete training run (15 trees, max depth 3), measured end-to-end over the real transport, client-side. The number in parentheses after each dataset name is the total number of records before the train/test split.

|  | **2-of-3** |  | **3-of-5** |  |
|---|---|---|---|---|
| **Dataset** | **Base** | **+Hiding** | **Base** | **+Hiding** |
| Heart Disease (297) | 769 | 1,305 | 1,106 | 1,997 |
| Breast Cancer (569) | 1,368 | 2,603 | 2,057 | 4,113 |
| Pima Diabetes (768) | 586 | 917 | 822 | 1,374 |
| CDC BRFSS (70,692) | 1,056 | 1,920 | 1,560 | 2,999 |

Path hiding increases traffic because each client submits histogram shares for every active node at a depth level. A fully expanded depth-three tree requires contributions for 1 + 2 + 4 = 7 nodes, compared with three node contributions without path hiding, giving a histogram-payload ratio of 7*/*3 ≈ 2.33. Actual overhead depends on the number of active nodes and is reduced in the total traffic by shared costs such as statistics collection and round-context downloads.

Increasing tree depth raises communication costs, particularly with path hiding, while predictive gains vary by dataset (Fig. 7). Pima Diabetes improves mainly up to depth three, CDC BRFSS continues to improve at greater depths, and Heart Disease and Breast Cancer show no consistent improvement across the tested depths. These results support depth three as a practical compromise between predictive performance and communication cost.

**Fig. 7:**
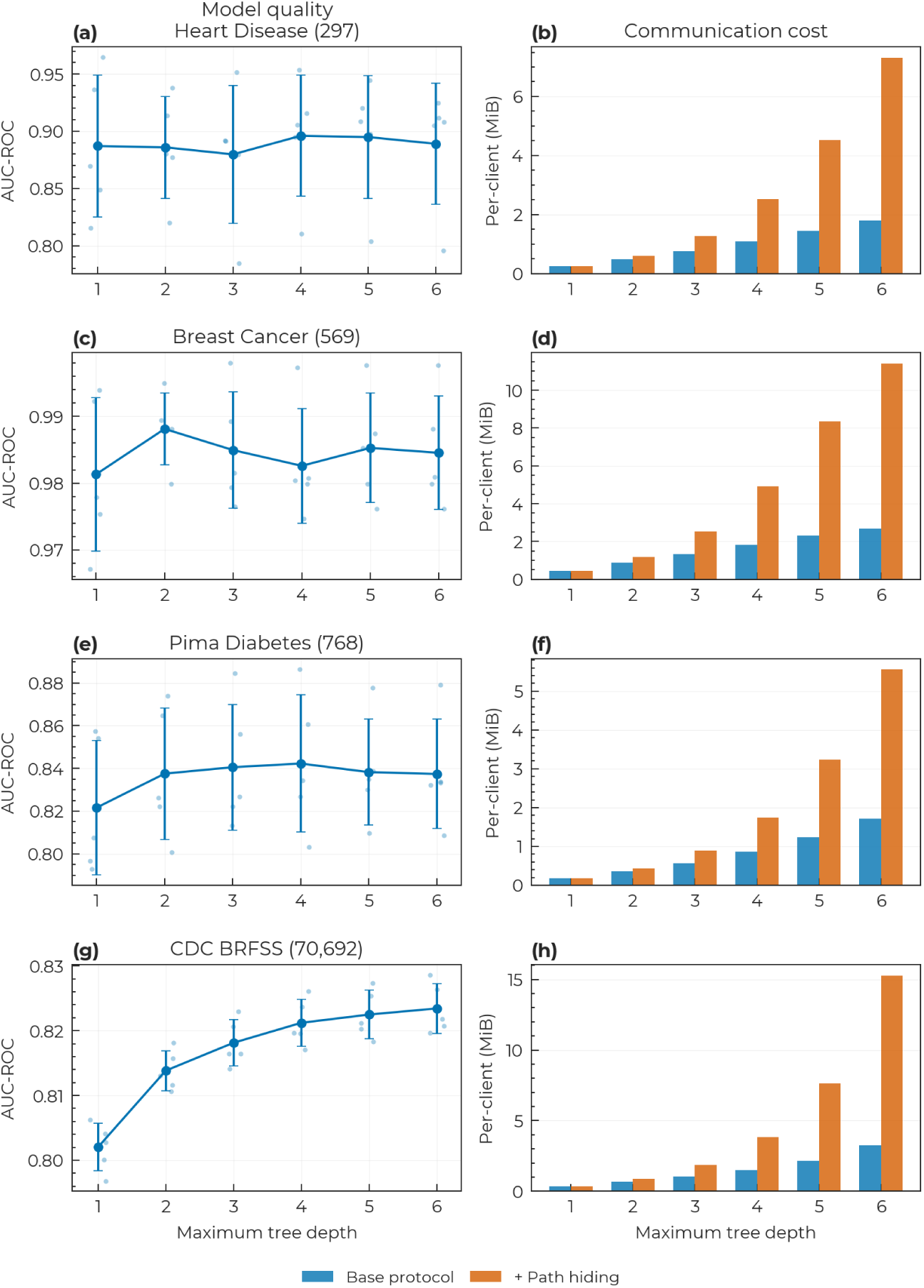
Model quality and communication cost as a function of tree depth. Panels are paired by dataset: Heart Disease (a, b), Breast Cancer (c, d), Pima Diabetes (e, f) and CDC BRFSS (g, h). Left column (a, c, e, g): AUC-ROC against maximum tree depth; data are mean ± s.d., translucent points show individual values (*n* = 5 independent stratified splits). Right column (b, d, f, h): measured per-client communication cost (MiB) with and without path hiding, one instrumented training run per configuration.

### Real-world deployment

We evaluated whether consumer smartphones and tablets could complete training through background execution without user interaction after enrolment. The deployment used operating-system-managed execution opportunities to compute and submit contributions while device screens remained off.

Clients used silent push notifications and scheduled background tasks to obtain execution opportunities. The aggregator sent at most one silent notification per device every 15 minutes while a training round was open. Clients also registered background tasks through WorkManager on Android and background app refresh on iOS. Execution depended on operating-system scheduling; the configured notification interval did not guarantee when a device would run.

An aggregator and three shareholders using a 2-of-3 Shamir threshold ran on one cloud server with 2 vCPUs and 4 GB RAM, with client connections protected by TLS. Four consumer devices—a Google Pixel 9 Pro, Samsung Galaxy Tab A8, Lenovo Tab M8 and Apple iPhone 13 Pro—each held one quarter of every training set. Heart Disease, Pima Diabetes and Breast Cancer sessions ran concurrently overnight, each training eight trees of maximum depth three. Submission windows lasted up to one hour, with each round closing once three devices had contributed. We recorded model quality, contribution timing and triggers, computation and submission times, resident memory and network traffic.

All three training sessions completed within six hours and seventeen minutes without a failed round (Figure 8). Final test AUC-ROC was 0.933 for Heart Disease, 0.821 for Pima Diabetes and 0.997 for Breast Cancer (Figure 9a).

**Fig. 8:**
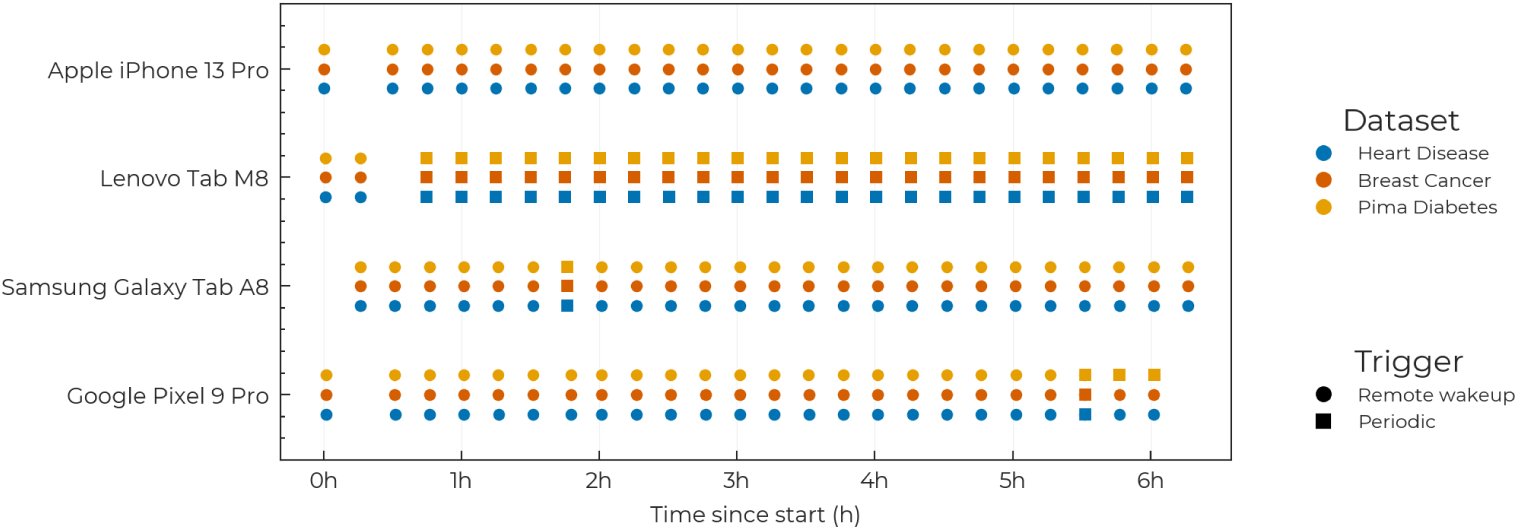
Timeline of the deployment. Each marker is one submitted round of the three concurrently trained sessions on the four devices; colour indicates the session, shape the wake trigger. Time is measured from the first submitted round of the capture.

**Fig. 9:**
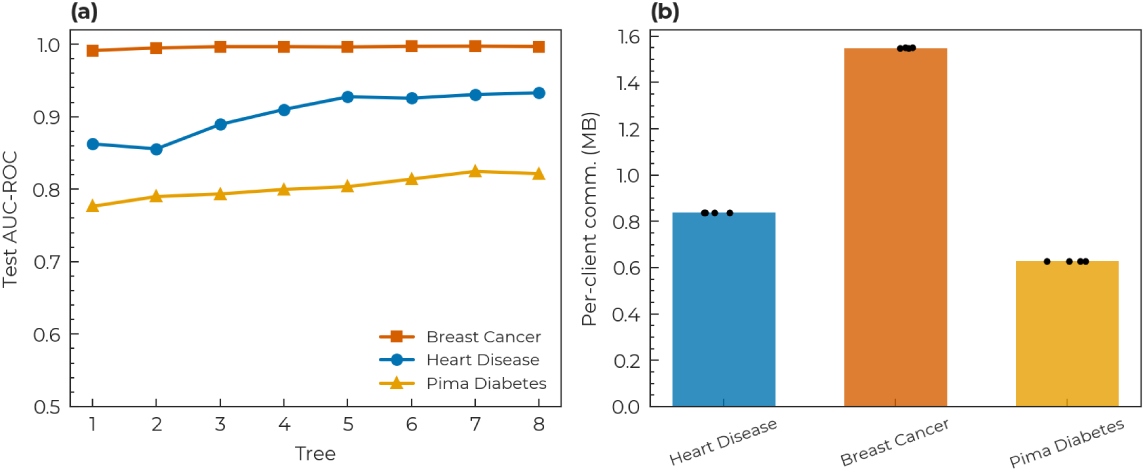
Model quality and per-client communication in the deployment. (a) Per-tree test AUC-ROC of the three concurrently trained sessions (8 trees of depth 3). (b) Measured per-client communication per session, summed over a device’s submitted rounds; bars show the mean, points the individual values (*n* = 4 devices).

Remote wakeups triggered 223 of the 300 device contributions, and scheduled background tasks triggered the remaining 77. The Lenovo tablet relied almost entirely on scheduled tasks (Figure 10a). On every device, the median interval between successive submitted rounds within a session was 15.0 minutes, matching the configured notification interval (Figure 10b).

**Fig. 10:**
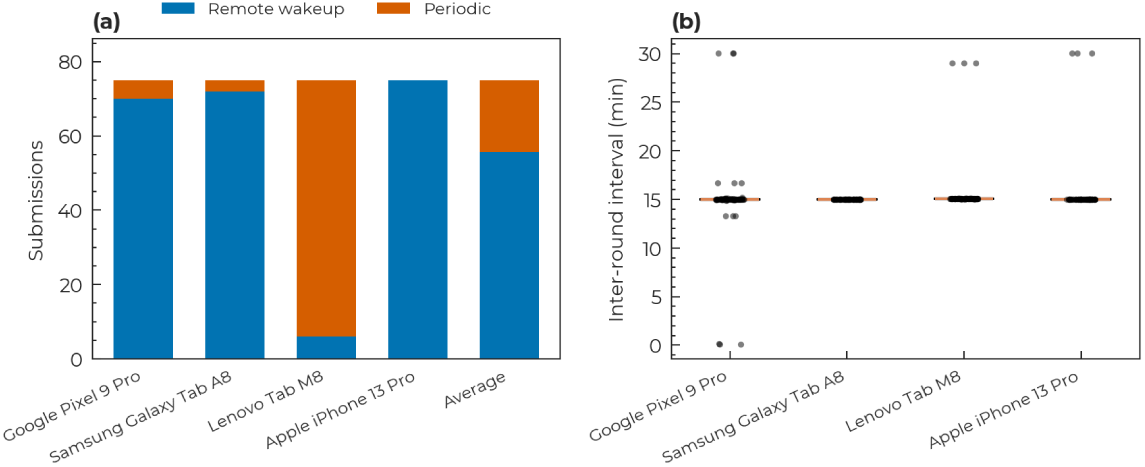
Participation in the deployment. (a) Submitted rounds per device by wake trigger, with the four-device average. (b) Minutes between a device’s successive submitted rounds within a session; boxes show the median and interquartile range, whiskers extend to 1.5× the interquartile range, and points show all individual intervals (*n* = 72 intervals per device).

Across devices, median share-generation times ranged from 0.3 to 1.0 milliseconds per round, and median submission times from 116 to 149 milliseconds over TLS.

Median application resident memory ranged from 76 to 234 MB (Table 7). Accumulated share-generation and submission time was 9.1–12.4 seconds per device across the three sessions. Measured per-client traffic was 0.63–1.55 MB per session (Figure 9b).

**Table 7:**
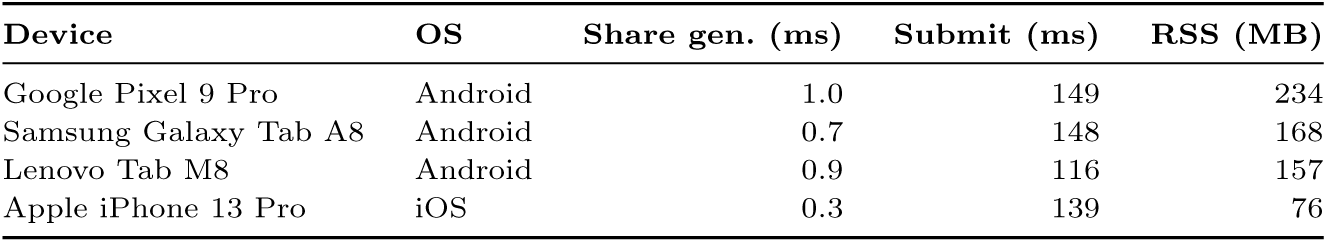
On-device cost of participating in the deployment. Perdevice medians over all submitted rounds (*n* = 75 rounds per device, across the three concurrent sessions). Share gen. is the time to compute one round’s secret shares over the device’s whole batch of records, Submit the time for the fan-out of those shares to the three shareholders over TLS, and RSS the resident set size of the application process.

| **Device** | **OS** | **Share gen. (ms)** | **Submit (ms)** | **RSS (MB)** |
|---|---|---|---|---|
| Google Pixel 9 Pro | Android | 1.0 | 149 | 234 |
| Samsung Galaxy Tab A8 | Android | 0.7 | 148 | 168 |
| Lenovo Tab M8 | Android | 0.9 | 116 | 157 |
| Apple iPhone 13 Pro | iOS | 0.3 | 139 | 76 |

The pilot demonstrates unattended training participation from consumer smartphones and tablets after one-tap enrolment per dataset. Computation and submission occupied only a small fraction of the elapsed training time, which was dominated by intervals between background execution opportunities. Battery consumption and perceived impact on device use were not measured.

## Discussion

PrivateBoost enables clients to contribute individual records to gradient-boosted tree training without fitting local models or coordinating with other clients. The simulations show predictive performance comparable to matched centralised training, while the physical pilot demonstrates unattended participation from consumer smartphones and tablets holding batches of records. In the pilot, computation and submission occupied only a small fraction of the elapsed training time. Under the evaluated notification and participation schedule, intervals between background execution opportunities dominated completion time, highlighting the importance of the interactions required from each client.

Clients can submit independently within a round, but training still follows the sequential dependencies of boosting. Each depth level depends on the preceding split decisions, and each tree uses predictions from the preceding ensemble. PrivateBoost accommodates changing participation between these rounds rather than requiring the same clients to remain available throughout training. The dropout experiments indicate that predictive performance can tolerate substantial randomly sampled nonparticipation in the evaluated datasets. In deployment, however, availability may correlate with device characteristics or participant behaviour, changing which records contribute and potentially introducing bias.

The use of external shareholders allows clients to finish their contribution to a round once their shares have been delivered. In Bonawitz et al.’s secure aggregation protocol, clients also participate in an unmasking phase after masked contributions have been collected. Individual clients may drop out after uploading, provided enough others supply the masking shares needed to complete aggregation [22]. These additional interactions may span multiple background execution windows. PrivateBoost can instead complete aggregation without further client responses once the selected shareholders hold a sufficiently large common set of contributions.

This design requires available shareholders and the stated limits on collusion. Client-based secure aggregation avoids the need for a fixed external shareholder set, and sparse-neighbourhood approaches reduce communication for coordination among clients [23]. With a fixed model configuration and shareholder set, PrivateBoost keeps each single-record client’s contribution workload independent of the client population, while infrastructure workload grows with participation. The four-device pilot establishes background participation with locally batched records and co-located server roles. Evaluating concurrent connections, polling and aggregation across independently operated shareholders would extend these findings to larger populations. Matched workloads and participation schedules would also allow the completion times of the two aggregation architectures to be compared.

The privacy guarantees concern both the numerical contributions and the information exposed by their submission. Threshold secret sharing protects individual contributions from coalitions of fewer than *m* shareholders, and path hiding conceals the node membership otherwise revealed by selective submissions. The aggregator nevertheless receives exact statistics, and the trained model also remains available for analysis. Gradient-reconstruction and membership-inference attacks illustrate how information about training records can be recovered from gradients or learned models [27, 28]. Although those studies examine neural-network settings, the distinction between protecting individual messages and protecting released information also applies here. Small node or bin populations can make aggregate outputs particularly revealing.

Differential privacy offers complementary protection by limiting the influence of an individual record on released information [37]. Maddock et al. demonstrate its application to federated boosted trees, including the trade-off between privacy and predictive utility [16]. PrivateBoost’s additive aggregation interface could support bounded contributions and calibrated noise, with privacy accounting covering both feature-statistics estimation and subsequent training releases. Protection against the aggregator would require perturbing aggregates before it observes them. Such an extension would need to account for repeated participation and evaluate the resulting effects on predictive performance.

Protecting against malicious clients requires additional checks before aggregation. Prio uses secret-shared non-interactive proofs to verify that submitted data satisfy specified validity constraints without revealing individual values [38]. Similar checks could enforce bounds on gradient and Hessian contributions and constraints on histogram structure, limiting manipulation through oversized or malformed submissions. Remote attestation provides a complementary basis for assessing client software integrity [39]. If contribution generation runs within an attested protected environment and its outputs are bound to the submitted shares, this could help enforce the prescribed computation. Neither approach establishes that the underlying records are truthful: deliberately misleading contributions within the permitted constraints would remain possible. These mechanisms are not part of the current implementation.

Infrastructure trust and participation metadata require separate consideration. The security analysis assumes honest-but-curious behaviour and excludes mixed coalitions, including collaboration between shareholders and the aggregator. Independent operation can support that separation, but does not enforce it cryptographically. Fresh round commitments also do not conceal network addresses, submission timing or persistent pseudonyms visible to shareholders. Anonymous communication could reduce some of this linkability, depending on the deployment. The designated shareholder organisations and their responsibilities can be specified before enrolment, while the applicable consent and data-governance arrangements depend on the deployment and jurisdiction.

Beyond the evaluated learner, the aggregation interface applies to training procedures expressed through additive per-record statistics. Histogram construction in LightGBM and the leaf-averaging step in CatBoost’s Plain-mode split evaluation provide related examples [40, 41], while formulations of first-order boosting also use additive statistics [42]. Supporting a complete learning algorithm requires examining its other operations: ordered categorical statistics, prefix-dependent ordered boosting, residual quantiles and selection based on individual gradient magnitudes are not supplied by the present interface. Tree-growth policies also change the balance between the number of aggregation rounds and the work performed in each round. These differences should guide extensions rather than assuming that compatibility with histogram aggregation establishes compatibility with the entire learner.

Tabular foundation models provide a further point of comparison. TabPFN achieved strong predictive performance on benchmarks with up to 10,000 samples and 500 features [43], but its published inference procedure uses individual labelled training examples as context. PrivateBoost’s aggregation interface exposes sums of per-record statistics, so it cannot directly support that procedure. Applying comparable privacy protection to prior-data fitted networks would require a different computational design [44].

The findings support a design in which personal devices perform brief local computations while infrastructure parties coordinate aggregation. The next evaluations should examine independently connected single-record clients under realistic participation patterns and measure the costs of stronger output privacy and contribution validation. Together, these would clarify the deployment settings in which the demonstrated mobile feasibility can translate into sustained training at larger scales.

## Conclusion

PrivateBoost enables federated gradient boosting from individual records without requiring clients to fit local models or coordinate with one another. Its fixedshareholder architecture combines secret-shared contributions with a client workload that remains independent of the participant population for a given model and shareholder configuration. Simulations across four medical datasets show predictive performance comparable to matched centralised training and resilience to randomly sampled client dropout.

The physical pilot demonstrates that consumer smartphones and tablets holding batches of records can complete training unattended through operating-systemmanaged background execution, with median share-generation times of 0.3–1.0 milliseconds per round. Together, these results establish a practical foundation for learning from records distributed across personal devices. By placing aggregation coordination on infrastructure parties, PrivateBoost allows participation to fit around intermittent device availability, bringing collaborative medical learning closer to the settings in which individuals hold and contribute their own data.

## Data Availability

All data produced in the present study are available upon reasonable request to the authors

## Acknowledgements

The authors would like to thank the Ministry of the Economy in Luxembourg and its digital health directorate for co-supporting this research. Similarly, we would like to thank the Luxembourg National Research Fund (FNR) for partially funding this research. This work was supported in part by a PhD grant from the Luxembourg National Research Fund (FNR) under the project reference 17223919/MMS/Industrial Fellowship, and in part by the R-MMS clinical trial.

## Author Contributions

B.S., S.G., and Z.T. conceived and designed the study. B.S. developed the methodology, curated the data, and implemented the software. B.S. and Z.T. performed the formal analysis. B.S. prepared the figures. B.S., O.E. and Z.T. wrote the original draft of the manuscript. Z.T. and S.G. administered the project. D.K., R.S., and Z.T. supervised the research. R.C. provided scientific advice. All authors contributed to reviewing and editing the manuscript. Following the passing of R.S. during peer review, the surviving authors reviewed and approved the final revised version.

## Data Availability

This study analysed four publicly available tabular health datasets. Three are held by the UCI Machine Learning Repository: Heart Disease [30, 31] (DOI 10.24432/C52P4X); Breast Cancer Wisconsin (Diagnostic) [32, 33] (DOI 10.24432/C5DW2B, also distributed with scikit-learn); and CDC Diabetes Health Indicators [36] (ID 891, DOI 10.24432/C53919), from which the class-balanced 70,692record subset analysed here is produced by the preparation script in the code repository. For Pima Indians Diabetes, the preparation script uses the CSV mirror maintained by Jason Brownlee [35]; the original study is described by Smith et al. [34]. No new patient data were collected: the real-world deployment partitioned these same public datasets across the participating devices. Deployment telemetry was collected in a cloud document database (Google Firestore) operated by the authors, and account and device identifiers were stripped before analysis. The datasets, training-run outputs and de-identified deployment records are publicly available with the analysis code on Zenodo (DOI 10.5281/zenodo.24354793), and figure source data are provided with this paper in the `Source_Data.xlsx workbook`.

## Code Availability

The source code for *privateboost* is publicly available at https://github.com/MyelinZ/ privateboost, released under the Apache 2.0 licence. The repository contains the protocol implementation, the server, client and mobile application, and the dataset preparation, experiment and figure-generation scripts. The implementation used in the experiments is archived as version 1.0.1 under DOI 10.5281/zenodo.21630278. The updated archive (version 1.0.2, DOI 10.5281/zenodo.24354793) includes the same implementation and experiment outputs, along with the revised analysis code [45].

